# Degradation of lipid based drug delivery formulations during nebulization

**DOI:** 10.1101/2021.03.16.21253714

**Authors:** David M. Klein, Albert Poortinga, Frank M. Verhoeven, Daniel Bonn, Sylvestre Bonnet, Cees J.M. van Rijn

## Abstract

Encapsulating pharmaceuticals in protective lipid based nanoparticles, and nebulizing them towards the target area in the body offers a range of clinical advantages. However, the process of nebulization might possibly damage sensitive nanoparticle structures, such as liposomes, resulting in loss of active pharmaceutical ingredients. We compare this loss for two types of lung inhalation devices: high-frequency piezo-actuated vibrating mesh nebulizers and non-actuated continuous jet nebulizers. We find that vibrating mesh nebulizers cause model liposomes to release more than ten times as much encapsulated material as the continuous jet nebulizers because the energies involved in nebulization are much larger. This result highlights the importance of applying a mild nebulization technology when administering shear-sensitive drug formulations such as lipid nanoparticle based drugs to the lungs.

## 1. Introduction

The nebulization route has some advantages over other administration routes for active pharmaceutical ingredients that specifically target the lung area. The intended therapeutic action in the lung can be triggered faster, unpleasant intravenous injections can be prevented, and also a smaller drug quantity is required, as the drug is delivered directly to its intended site of action. This mode of administration typically results in lower systemic side-effects in comparison to oral or intravenous administration. Side-effects can be further suppressed by encapsulating the drug inside liposomes or lipid nanoparticles (LNPs) [1,2]. Indeed, entrapping drugs in ‘conventional’ liposomes has proven advantages such as targeted drug delivery to specific tissues, and prevention against drug degradation [1]. Such protection is appealing for drug and vaccine makers [3,4], for example in the case of RNA drugs which are vulnerable to RNase degradation [3,5]. In addition, the use of LNPs facilitates cellular uptake of the drug with a high efficacy [2]. LNPs can be seen as a new generation of liposomes, specifically formulated for an efficient delivery of various active pharmaceutical ingredients, and are characterized by having a smaller internal aqueous content than liposomes [1]. However, in the process of converting the lipid based nanoparticle drug formulations into aerosol droplets (nebulization), the nanoparticles may get damaged due to shear degradation, resulting in loss of the originally entrapped active pharmaceutical ingredients, in particular in case of hydrophilic materials [6]. This is due to a high shear stress being exerted on the nanoparticles, leading to breakage. The development of lipid based nanoparticle carriers for inhalation is therefore focused on increasing the strength and rigidity of the nanoparticles with the aim of reducing the detrimental effect of shear stress on nanoparticle stability and maximizing its deposition rate and efficacy of the formulation in the ‘deep lung’. An example of a lipid based nanoparticle drug formulation success is the recently FDA approved nebulizable liposome formulation of the antibiotic Amikacin, the success of which is attributed to the combined development of a shear-stress resistant nanoparticle formulation of cholesterol-enriched dipalmitoyl-phosphatidylcholine (DPPC-CH) with a liposome size around 300 nm and a PARI eFlow vibrating mesh nebulization device [7]. In this communication we investigate a novel facile and fast nebulization method with a continuous Rayleigh jet atomization device to nebulize formulations at a throughput well over 1 mL/min with a minimum amount of shear stress enabling a large window to formulate lipid based nanoparticle drug carriers.

## 2. Materials and methods

### Preparation of DPPC liposome formulations

The osmolarity of aqueous solutions was measured on a Micro-Osmometer Autocal Type 13 from Roebling. Calcein was obtained from Carl Roth and used as received. 1,2-dipalmitoyl-sn-glycero-3-phosphocholine (DPPC) and cholesterol (CH) were purchased from Avanti Polar Lipids and sodium N-(carboyl-methoxypolyethylene glycol-2000)-1,2-distearoyl-sn-glycero-3-phosphoethanol-amine (DSPEPEG2K) from Lipoid. All were stored as solids at −20 °C. Liposomes were prepared as follows [8,9]. The lipids were dissolved in chloroform at the desired ratio (DPPC 100, DPPC-CH 50:50, and DPPC-DSPEPEG2K 100:1) in a pressure resistant glass tube. Chloroform was evaporated by rotary evaporation and the resulting lipid film was dried in vacuum overnight to remove residual solvent. The film was then hydrated with a NaH_2_PO_4_ buffer (1 mL, 0.1 M, pH = 7.7, p = 661 mOsm) containing calcein (70 mM), followed by 5 freeze-thaw cycles between liquid N_2_ and a 50 °C water bath. Subsequently, the vesicles were extruded 11 times with an Avanti Polar Lipids mini-extruder through a 100 nm polycarbonate membrane at 55 °C. After extrusion, the liposomes were separated from the non-encapsulated calcein using a SEC column (GE Healthcare cartridge) equilibrated with NaH_2_PO_4_ buffer (0.1 M, pH = 7.7, p = 663 mOsm, osmolarity adjusted by adding NaCl). The orange/brown non-fluorescence band containing the liposomes was obtained until free calcein eluted, as visualized by UV light. These liposome stock solutions (final bulk lipid concentration 5 mM, assuming no losses) were analyzed the same day with dynamic light scattering (DLS) and calcein luminescence. DLS was performed at 25 °C on a Zetasizer Nano-S from Malvern operating at 632.8 nm with a scattering angle of 173 °C.

### Calcein leakage testing

To a 1 mL cuvette was added 0.04 mL liposome stock solution and an isotonic 0.96 mL NaH_2_PO_4_ buffer (0.1 M, pH = 7.7, p = 663 mOsm). A luminescence measurement was carried out on a Horriba Aqualog spectrometer at RT using 495 nm as excitation wavelength. The emission intensity of calcein was recorded at 519 nm. The maximum luminescence intensity of calcein at 519 nm of the sample was determined by addition of TritonX100 (10 mM, 5 µL, 16 days equilibration time), which resulted in the destruction of the liposomes and subsequent release of all calcein into the bulk aqueous solution. The percentage of release of encapsulated calcein (%) was calculated by dividing the fluorescence intensity at a given time, by the final, maximum fluorescence intensity obtained after TritonX100 addition.

### Nebulizers

The nebulizers used in this study are shown in Fig.1. Nebulizer A has a flow rate of 0.4 mL/min, nebulizer B of 0.5 mL/min and nebulizer C of 1.5 mL/min. Nebulizer A and nebulizer B were filled with 2 mL of liposome formulation and Nebulizer C with 1 mL of liposome formulation. 1 mL of the formulation was nebulized. The nebulized formulation was collected by spraying into a 50 mL centrifuge tube. The degree of calcein leakage inside the collected fluid was measured as described above. The 1 mL liposome formulation that remained in the reservoir of nebulizer A and B at the end of each nebulization experiment was also analyzed for calcein release. Nebulization was undertaken within 2 h of preparation of the liposome formulation.

**Figure 1.**
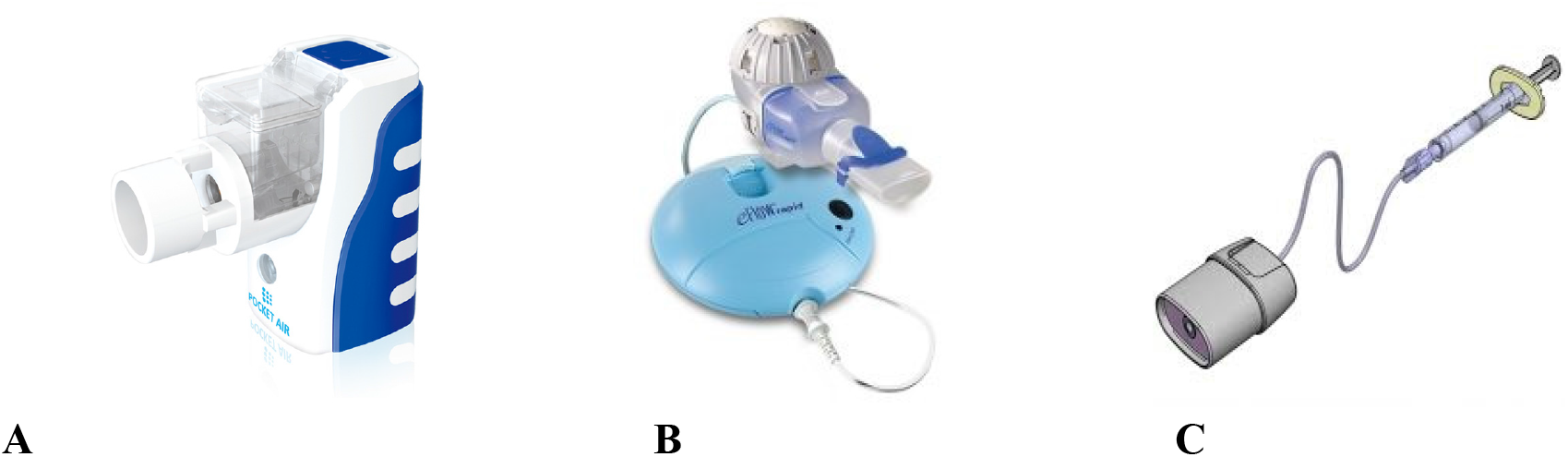
Nebulizers used in this study. (A) Pocket air, vibrating mesh nebulizer, (B) PARI eFlow, vibrating mesh nebulizer, (C) Continuous Rayleigh jet nebulizer.

### Estimating the energy dissipation of the actuated nebulizers

To estimate the dissipation of the actuated mesh nebulizers water was sprayed for 1 min and the temperature increase of the mesh and reservoir was measured using a Flir C3 infrared camera. The measured temperature increase per second was multiplied with the heat capacity of water and divided by the flow rate to arrive at a value for the so-called energy density (in J/g): the amount of energy added to the sprayed product.

## 3. Results and Discussion

To ensure that the results apply to different lipid formulations, we prepared three different types of liposomes, i.e. DPPC, DPPC-CH 50:50, and DPPC-DSPEPEG2K 100:1, encapsulating calcein in their inner aqueous compartment as a model of a negatively charged drug. Calcein can be used for testing membrane leakage [10]; it is a self-quenching fluorophore that shows low fluorescence at high concentration inside the liposome (70 mM), but increased fluorescence at lower concentration, for example when it leaks outside the liposome into the bulk. Destroying the liposome membrane by adding the surfactant TritonX100 afforded a maximum fluorescence intensity F_max_, which allowed to quantify the relative fraction of drug released before full membrane disruption, X (in %), by dividing the fluorescence intensity F (before or after nebulization) by F_max_. Figure 2 depicts the hydrodynamic size distributions of the calcein-encapsulating DPPC, DPPC-CH and DPPC-DSPEPEG2K liposome formulations in phosphate buffer 1 h after preparation, as determined by DLS. Significant aggregation of the pure DPPC (Fig. 2A) and DPPC-CH [1:1] formulations (Fig. 2B) was observed, whereas the 1% PEGylated DPPC formulation (stealth liposomes) did not aggregate at all, as characterized by a low polydispersity index (PDI < 0.1, Fig. 2C). The characterization of these size distributions is given in Table 1.

**Table 1.**
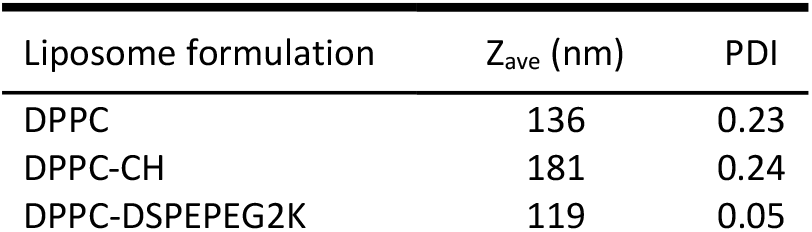
Characterization of the liposomes by DLS

| Liposome formulation | $Z_{ave}$ (nm) | PDI |
| --- | --- | --- |
| DPPC | 136 | 0.23 |
| DPPC-CH | 181 | 0.24 |
| DPPC-DSPEPEG2K | 119 | 0.05 |

**Figure 2.**
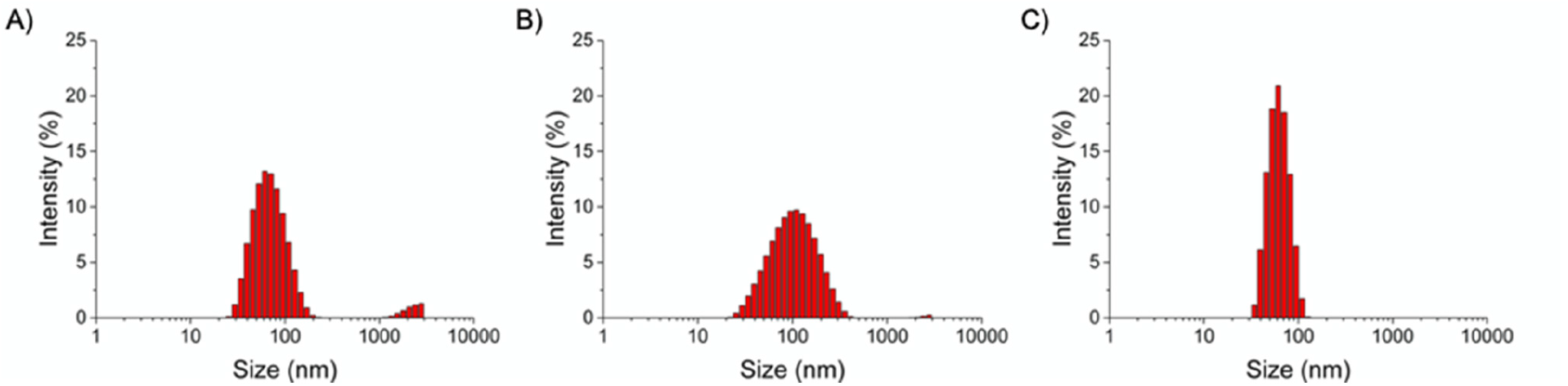
Hydrodynamic size distributions of calcein-encapsulating liposome formulations (A) DPPC, (B) DPPC-CH 50:50, and (C) DPPC-DSPEPEG2K 100:1.

In a second step, the relative calcein leakage of each formulation was quantified by measuring the fluorescence intensity F at the maximum emission (519 nm) of calcein, either directly after preparation, 1 h after preparation, or after nebulization, and dividing it by the maximum emission intensity F_max_ when 100% of calcein was released by TritonX100-induced destruction of the liposome membrane. While storage of the liposomes for 1 h did not lead to measurable calcein leakage (variation <0.1%), all liposome formulations released calcein after nebulization (Table 2). However, clear differences in calcein leakage were observed, depending on the type of nebulizer. The actuated mesh nebulizers caused a lot of calcein leakage outside the liposome (13-16% with the pocket air and 32-37% with the Pari eflow), while the continuous Rayleigh jet nebulizer caused negligible drug leakage (2.3-4.6%). These results confirmed that it is the nebulization procedure that induced drug release outside the liposome, and not thermal leakage of the membrane. Secondly, they highlight the difficulty of nebulizing drug-encapsulating liposomes for example for delivery to the lungs: the energy used to generate the droplets locally tears the lipid membrane, thus leading to unwanted drug release before the liposomes have reached their target. Another potential issue is the heat generated in the reservoir by vibrating mesh nebulizer, which as observed here may also lead to up to 24% of calcein leakage. Thermal effects on membrane leakage have been reviewed recently [11].

**Table 2.** Calcein release after nebulization

| Experiment<sup>a</sup> | Liposome formulation |  |  |
| --- | --- | --- | --- |
|  | DPPC | DPPC-CH | DPPC-DSPEPEG2K |
| Pocket air mist | 15 ± 1% | 13 ± 1% | 16 ± 3% |
| Pocket air reservoir | 14 ± 1% | 13 ± 3% | 4.8 ± 0.9% |
| Pari eflow mist | 37 ± 2% | 32 ± 1% | 34 ± 9% |
| Pari eflow reservoir | 24 ± 1% | 23 ± <1% | 6.6 ± 2.1% |
| Rayleigh jet mist | 4.6 ± 0.7% | 2.3 ± 0.3% | 2.6 ± 2.6% |

<sup>a</sup> all experiments were performed in duplicate.

In order to explain the high leakage observed during nebulization using mesh nebulizers we determined the energy density dissipated in the solution during nebulization. The energy density is a parameter that is commonly used to compare emulsification effects [12]. For the non-actuated mesh nebulizer, the energy density is proportional to the applied pressure, which is estimated to be 20 bar giving an energy density of 2 J/g. For the actuated vibrating mesh nebulizers, we calculated the energy density to be in the order of 30-60 J/g based on the temperature rise (>2 °C per 30 seconds for the Pocket air and >4 °C per 30 seconds for the Pari eflow) of the liquid in the reservoir after nebulization. In the actuated-mesh nebulizers droplets are produced by the action of high frequency pressure waves with a frequency in the range of 100 kHz. On the other hand the use of high frequency ultrasound waves is also a well-known technique to break or disrupt liposomes [13]. It has been reported that the decrease in liposome size is proportional to the energy density [14]. Using a frequency in the order of 100 kHz it was found that the average liposome size decreased with about ten percent at an energy density of 50 J/g. This makes it conceivable that pressure waves used in the actuated-mesh nebulizers cause the liposome membrane to deform and potentially disrupt, leading to a substantial loss of encapsulated calcein. Interestingly this assumption is now experimentally verified, as indeed a substantial leakage of calcein was found in the reservoir of the vibrating mesh nebulizers after nebulization (5-14% with the pocket air and 7-24% with the Pari eflow, see Table 2). In the syringe ‘reservoir’ of the Rayleigh jet nebulizer as expected no loss of the liposomes has been found.

### Liposomes and mechanical rigidity

DPPC liposomes are characterized by hydrogen saturated acyl chains, a high gel-to-liquid phase transition temperature (41.4 °C), and hence a high packing density at room temperature. Such characteristics lead to a gel-like, mechanically rigid, and stress-insensitive liposomes. Indeed, in absence of nebulization none of the formulation used here leaked significant calcein within 1 h. It is generally accepted that adding cholesterol (CH) make lipid membranes based on saturated acyl chains (e.g., DPPC) more fluid and lipid packing tighter, resulting in less leaky membranes. The addition of PEGylated phospholipids slightly has been reported to weaken liposomes, because PEGylated phospholipids have a higher exchange rate with the aqueous phase [15]. In absence of nebulization, none of these additives led to significant changes of the membrane leakage, as pure DPPC membranes were found already very tight at room temperature (<0.1% leakage within 1 h). Upon “hard” nebulization using vibrating mesh nebulizer, adding cholesterol to the DPPC formulation had minimal effects on reducing calcein leakage, but adding PEGylated lipids did increase the amount of calcein release, especially when passing the pores of the vibrating mesh, considering that in the reservoirs of the vibrating mesh nebulizers the PEGylated lipids remained well preserved during nebulization. This effect is reminiscent from the effect of air bubbles on the leakage of 1,2-dioleoyl-sn-glycero-3-phosphocholine (DOPC) liposomes, which was increased in presence of PEGylated lipids in the membrane [16]. Upon “soft” nebulization using Rayleigh jet nebulizer, both additives slightly reduced calcein release, which remained much lower than when nebulization was performed using the vibrating mesh nebulizers. In conclusion, we have put forward an efficacious Rayleigh jet inhalation technology that causes less unwanted leakage of encapsulated drugs outside the liposomes during nebulization, and this is regardless of the liposome composition. This nebulizing technology therefore enables nebulizing liposomes with a minimal escape of hydrophilic or large encapsulated drug molecules, such as DNA, RNA, proteins, peptides, antibodies, etc. while offering improved freedom in the formulation of lipid drug delivery systems.

## Data Availability

All data are available on request by corresponding author

